# Genetic links in angioimmunoblastic T-cell lymphoma (AITL), clonal hematopoiesis and concomitant hematologic malignancies provide insights into the cell of origin, etiology and biomarker discovery for AITL

**DOI:** 10.1101/2020.11.25.20238220

**Authors:** Shuhua Cheng, Wei Zhang, Giorgio Inghirami, Wayne Tam

## Abstract

We generated and compared the mutation profiles through targeted sequencing of the primary tumors and matched bone marrow/peripheral blood samples in 25 patients with angioimmunoblastic T-cell lymphoma (AITL) and 2 with peripheral T-cell lymphoma, not otherwise specified (PTCL-NOS). Our results provided strong evidence that AITL/PTCL-NOS, clonal hematopoiesis (CH) as well as other concomitant myeloid and even B-cell hematologic neoplasms (CHN), frequently arose from common mutated hematopoietic stem cell clones. Aberrant AID/APOBEC activity-associated substitutions and tobacco smoking-associated substitutions were enriched in the early CH-associated mutations and late non-CH associated mutations during AITL/PTCL-NOS development, respectively. Moreover, survival analysis showed that the presence of CH harboring ≥ 2 pathogenic TET2 variants with ≥ 15% of allele burden conferred higher risk for CHN (P = 0.0034, hazard ratio = 10.81). These findings provide insights into the cell origin and etiology of AITL, and provide a novel stratification biomarker for CHN risk in AITL/PTCL-NOS patients.

## Introduction

Peripheral T cell lymphoma (PTCL) is a heterogenous group of lymphoid tumors and encompass PTCL-NOS, angioimmunoblastic T cell lymphoma (AITL) and several other entities of T-cell lymphoma ^1^, likely driven by an array of recurrent genomic defects^2^. Except for PTCL-NOS, AITL is the most common subtype of PTCL (18.5% of mature T cell lymphoma) and is believed to arise from a subset of peripheral mature CD4+ T cells corresponding to follicular helper T (TFH) cells, characterized immunophenotypically by expression of a set of cellular markers like PD1, CXCR5, BCL-6, CD10, CXCL13 and ICOS-1 ^3-7^. Morphologically, AITL is typically characterized by a polymorphous lymphoid infiltrate with a proliferation of medium-sized tumor cells with clear cytoplasm (clear cell immunoblasts), associated with prominent proliferation of high endothelial venules and follicular dendritic cells. A subset of PTCL, termed as PTCL with TFH phenotype in the updated WHO classification, may be biologically related to AITL, sharing some clinic-pathologic features with AITL and is thought to be also derived from TFH 8. Although progress has been made in understanding AITL pathogenesis and developing new treatment^9^, AITL remains as an aggressive lymphoid tumor, with low estimated rates of overall and failure-free survival at five years (33% and 18%, respectively) ^10^. To develop more effective therapeutic agents against AITL and PTCL in general, with TFH phenotype further understanding of the molecular pathogenic mechanisms of AITL is needed.

Genetically, AITL is characterized by a number of genomic mutations in *TET2, RHOA, DNMT3A* and *IDH2* ^2,11-14^. Cell-intrinsic and/or extrinsic factors that facilitate the accumulation of these AITL-related mutations remain unclear. Mutations in *TET2* and *DNMT3A* are also frequently associated with myeloid malignancies, including acute myeloid leukemia (AML), myeloproliferative neoplasms (MPN) and myelodysplastic/myeloproliferative neoplasms (MDS/MPN). *TET2* and *DNMT3A* are also the most commonly mutated genes associated with clonal hematopoiesis (CH) in healthy adults, especially those over 60 years of age. CH has been shown to be an aging-related process characterized by the clonal expansion of hematopoietic cells harboring one or more somatic mutations, as a result of selective advantage in the hematopoietic stem and progenitor cells due to enhanced self-renewal and inhibition of differentiation ^15-18^. It has been noted that myeloid and lymphoid malignancies may co-occur in the same patients ^19^. Considering the similarities in genomic mutation profiles of AITL, myeloid malignancies and CH, it has been postulated that there may be a biological link between these entities. To test this, the current study implemented next generation sequencing (NGS) approach to analyze neoplastic T cells and paired bone marrow/peripheral blood specimens from a cohort of 27 patients with AITL or PTCL-NOS, and explored the potential of using the genomic findings from this study to shed light into the etiology of AITL development and to predict clinical progression related to development of second hematologic neoplasms, which is one of the clinical challenges in the clinical management of these cancer patients.

## Results

### Mutation profiling of AITL/PTCL-NOS and matched BM/PB supports an origin of AITL/PTCL-NOS from mutated HSCs associated with clonal hematopoiesis

For mutation profiling, we sequenced 27 pairs of AITL or PTCL-NOS samples using a 538-gene targeted NGS panel that covered recurrently mutated genes associated with T-cell lymphomas ^20^. Of the genomic regions targeted by the panel, 90% had a coverage depth of >1,000. Those sequenced samples included 27 diagnostic lymph node (LN) specimens from patients with AITL (n = 25) or PTCL-NOS (n = 2) and their corresponding bone marrow (BM, n=23) or peripheral blood samples (PB, n =4) from our archived specimens (hereafter denoted as AITL/PTCL-NOS). The overall genomic and pathologic findings showed that of the 27 BM/PB samples, 10 had no detectable involvement by AITL or PTCL-NOS (37%), while 17 were involved by the neoplastic T cells (63%) of variable abundance. One BM sample showed concomitant diagnostic involvement by a myeloproliferative neoplasm (MPN) (Patient #20) (**Supplementary Table1** and **Supplementary Fig.1**).

The genomic alterations found in the matched BM/PB can be due to (i) BM/PB involvement by AITL/PTCL-NOS; (ii) CH; or both (i) and (ii). To accurately characterize the mutation spectrum in the BM/PB, we distinguished the CH-associated mutations from those attributed to the BM/PB involvement by the T neoplastic cells according to the following algorithm. The tumor burden (TB) was estimated for each of the BM/PB specimens involved by the lymphomas (16 AITL and 1 PTCL-NOS) based on their histologic, immunophenotyping and T-cell receptor gamma (*TCRG*) gene rearrangement findings (**Supplementary Table1, Supplementary Fig.1C**), and compared to the variant allele frequencies (VAFs) of the somatic alterations of T-cell lymphoma-associated genes, for example *RHOA* p. G17V, a molecular characteristic of AITL ^11,12,14,21^. The AITL/PTCL-NOS-related variants present in the BM/PBs are highlighted with rectangles on the heat map (**Supplementary Fig.1C**) or colored in red in **Supplementary Table2**, and their VAFs (VAF^inv^) were found to be 1.06% on average (median, 0.565%; range: 0.1 - 5.64%) (**Supplementary Fig.1D**). The variants whose VAFs could not be attributed to AITL/PTCL-NOS involvement alone in the BM/PB, or the variants detected in BM/PB uninvolved by lymphoma were presumed to correspond to variants related to CH, which was confirmed by the presence of these CH-associated mutations in purified neutrophils in the peripheral blood of one patient (Patient #24, **Supplementary Table2** and **Supplementary Fig.2**). Compared to the AITL/PTCL-NOS-related variants, the VAFs of the CH-associated variants (VAF^CH^) were significantly higher (p<0.0001), ranging from 0.24% to 51.5% with a mean of 22.5% (median: 16.5%), approximately 21.2 times higher than the VAF^inv^ on average (**Supplementary Fig.1 D**).

We identified in the matched BM/PB specimens 44 variants from 14 genes, excluding the variants attributed to the BM/PB involvement by AITL/PTCL-NOS as described above (**Supplementary Fig.1A**). These alterations included 17 missense (38.64%) and 12 nonsense (27.27%) SNVs, 6 frameshift (13.64%) deletions, 7 frameshift (15.91%) and 1 in-frame insertions (**Fig. 1, Supplementary Fig. 3, Supplementary Table3**). Among these 44 somatic mutations, 37 mutations, identified in 19 of the 27 (70.4%) cases, were shared with those found in the primary lymphoma, and seven were BM/PB specific (**Fig. 1B, 1C, 1E, Supplementary Fig. 4**). The recurrent shared mutations were primarily restricted to *TET2* (74% of the cases) and *DNMT3A* (37% of the cases), consistent with the top CH-associated genes previously reported (**Fig. 1B, 1C**) ^22^. All but two (Pt #5, #20) of the cases with CH-associated mutations in the BM/PB did not have a dx of an overt myeloid neoplasm. These CH-associated mutations presumably were acquired very early in the common ancestral hematopoietic stem cells (HSC) from which both the myeloid and T-cell lineages are derived. Consequently, we defined these shared CH-associated variants as early mutations as seen below. In 8 patients (Patient #7, #9, #11, #12, #17, #21, #23, #26) (29.6%), no CH-associated mutations were detected in the BM or PB.

**Fig. 1.**
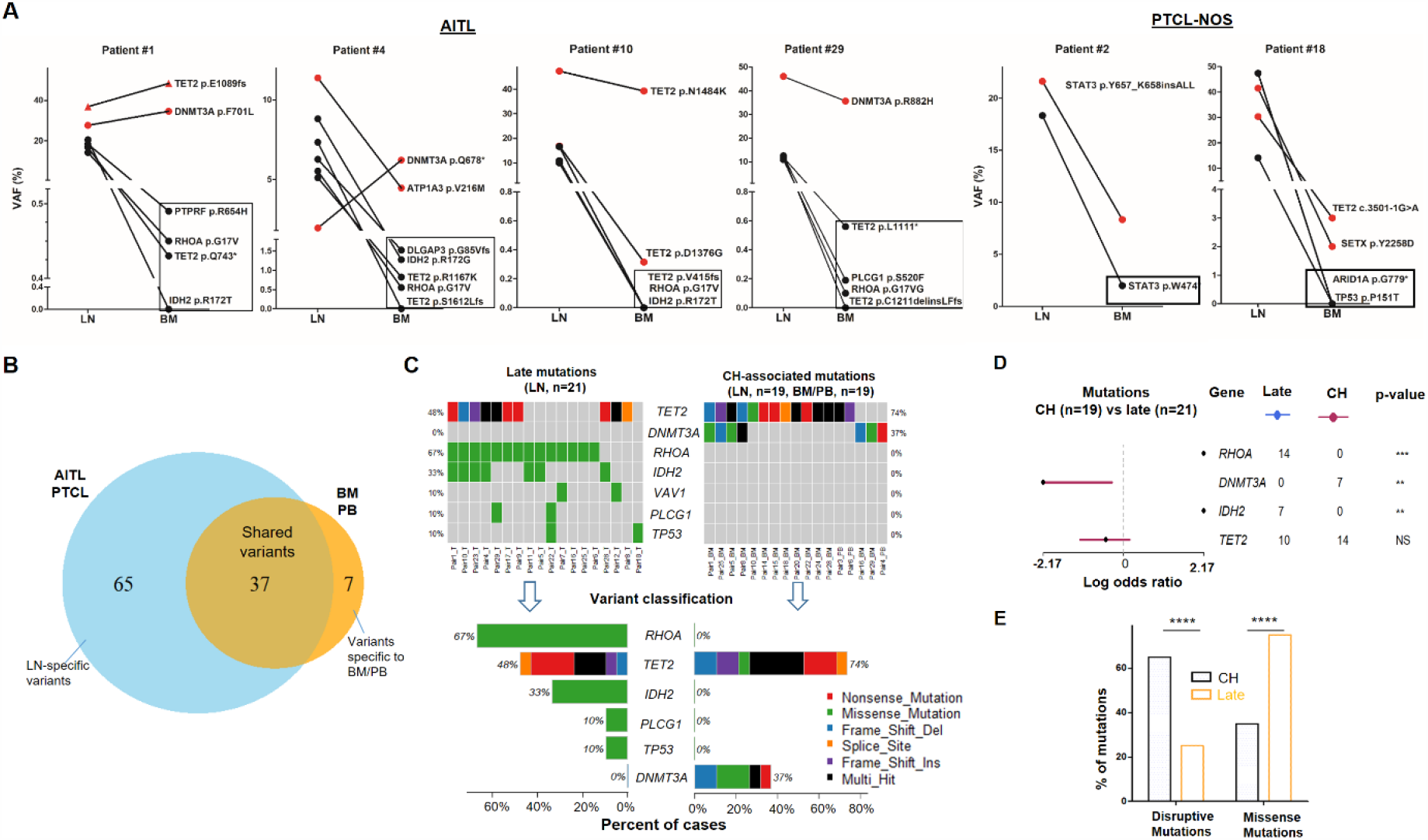
Analysis of genomic alterations by target sequencing panel for primary lymphomas and paired BM/PB in patients with AITL and PTCL-NOS. (A) Presence of CH in patients with AITL and PTCL-NOS. Dot plots showing the detected variants and their VAFs in the AITL and PTCL-NOS (LN) and their matched BM/PB in representative AITL and PTCL-NOS cases with CH. The black circles indicate variants specific to the lymphomas, and the variants shared between the primary lymphomas and CH are highlighted in red. The variants attributed to lymphoma only are boxed. VAF, variant allele frequency; LN, lymph node; BM, bone marrow; PB, peripheral blood; CH, clonal hematopoiesis. Additional detailed descriptions of these illustrative cases are provided in **Supplementary Text**. (B) Venn diagram showing the distribution of the shared, lymphoma or BM/PB-specific variants identified in the diagnostic LN and paired BM/PB samples. The shared variants are defined as variants identified in both the primary lymphoma and the BM/PB, the latter as CH-related variants. The variants predicted to be due only to lymphoma involvement in BM/PB have been excluded (see also **Supplementary Fig.1** for the distribution of all variants). (C) Summary of the CH-associated mutations in the BM/PB and LN, and the mutations postulated to accumulate at a later stage of lymphoma development (late mutations). The CH-associated mutations are shared between the primary lymphomas and the BM/PB and can be considered as early lesions in AITL/PTCL. The heat maps show the top recurrent mutations in both categories. Stacked bar plots show the type of variants and the mutation frequency (relative to our cohort) for each of the major mutated genes in the LN and BM/PB samples. (D) Forest plot showing differentially affected genes between the CH-associated and late mutations. (E) Comparison of the distribution of disruptive and missense mutations in the CH-associated and late mutations. * p < 0.05; ** p < 0.01; *** p < 0.001; **** p < 0.0001; NS, not significant.

In the 27 diagnostic LN samples, we identified a total of 102 non-synonymous somatic mutations in 37 genes with a median of ∼3 variants per sample, including 62 missense (60.78%) and 20 nonsense (19.61%) single-nucleotide variants (SNVs), 1 in-frame (0.98%) and 9 frameshift (8.8%) deletions, 1 in-frame and 7 frameshift (6.9%) insertions (**Fig. 1B, Supplementary Table3, Supplementary Fig. 5**). Of these 102 mutations, 37 were associated and shared with CH (**Fig. 1B, 1C, 1E**). In more than half of the T lymphoma cases, not only could we detect early CH-associated mutations, we also identified 65 mutations that are likely acquired during the later stage of AITL/PTCL-NOS development (referred as late mutations hereinafter) (**Fig.1B, 1C, Supplementary Fig. 4**). The recurrent late mutations were limited to several oncogenes and tumor suppressor genes, including the well-known driver genes like *RHOA* (67% of the cases), *TET2* (48%), *IDH2* (33%), *PLCG1*(10%), *TP53*(10%), *VAV1* (10%), and are characterized both by the absence of *DNMT3A* mutations (**Fig. 1C, 1D**) and by the enrichment of missense mutations, which were increased from 36.1% in the CH-associated mutations to 75.2% in the late mutations (proportion test, P value < 0.0001) (**Fig. 1C** and **1E**). The mutations in *IDH2, PLCG1, TP53*, were found exclusively as late mutations and not CH-associated mutations (**Fig. 1C, 1D**).

**Fig.1A** shows 4 representative AITL cases and 2 PTCL-NOS cases with their matched BM/PB, where red dots indicate the CH-associated variants present in both the primary lymphoma and BM/PB, and black dots represent the variants associated with AITL/PTCL-NOS (also highlighted with rectangles). Detailed description of these illustrative cases is provided in **Supplemental Text**. There are a couple of notable findings: first, more than one CH clone can be present in the BM, and their clonal representations in the BM may not reflect those in the lymphoma, as seen in the *DNMT3A*-mutated clones in patient #4. These results suggest that the same *DNMT3A* mutation can have differential effect depending on the cell lineage affected. Second, findings in patient #29 raise the possibility that besides the neoplastic T cells, reactive lymphocytes in these two cases might also harbor the CH-associated mutations.

Our results support a tumor model in which AITL/PTCL, NOS emerges from mutated and expanded HSC clones that are associated with CH in the BM as well as serving as the lymphoma precursors. The latter often acquires additional missense mutations during the course of development to frank lymphomas.

### Late mutations in AITL/PTCL, NOS are enriched for C>A transversion substitutions possibly associated with smoking

We investigated whether there might be an etiologic difference between the CH-related mutations and the late mutations by analyzing the type of substitutions for these mutations.

For the CH-associated mutations, overall transition (Ti) and transverse (Tv) substitution rates are comparable (Ti vs Tv, median, 50% vs 50%, mean, 54.76% vs 45.24%, P value >0.05) (**Fig. 2A**). At the base substitution level, C>T is found most frequently (44%), followed by C>G (20%) (**Fig. 2A**), which is consistent with the reported aging and/or overactivity of APOBEC family members-associated mutational process through replicative mutagenesis ^23^.

**Fig. 2.**
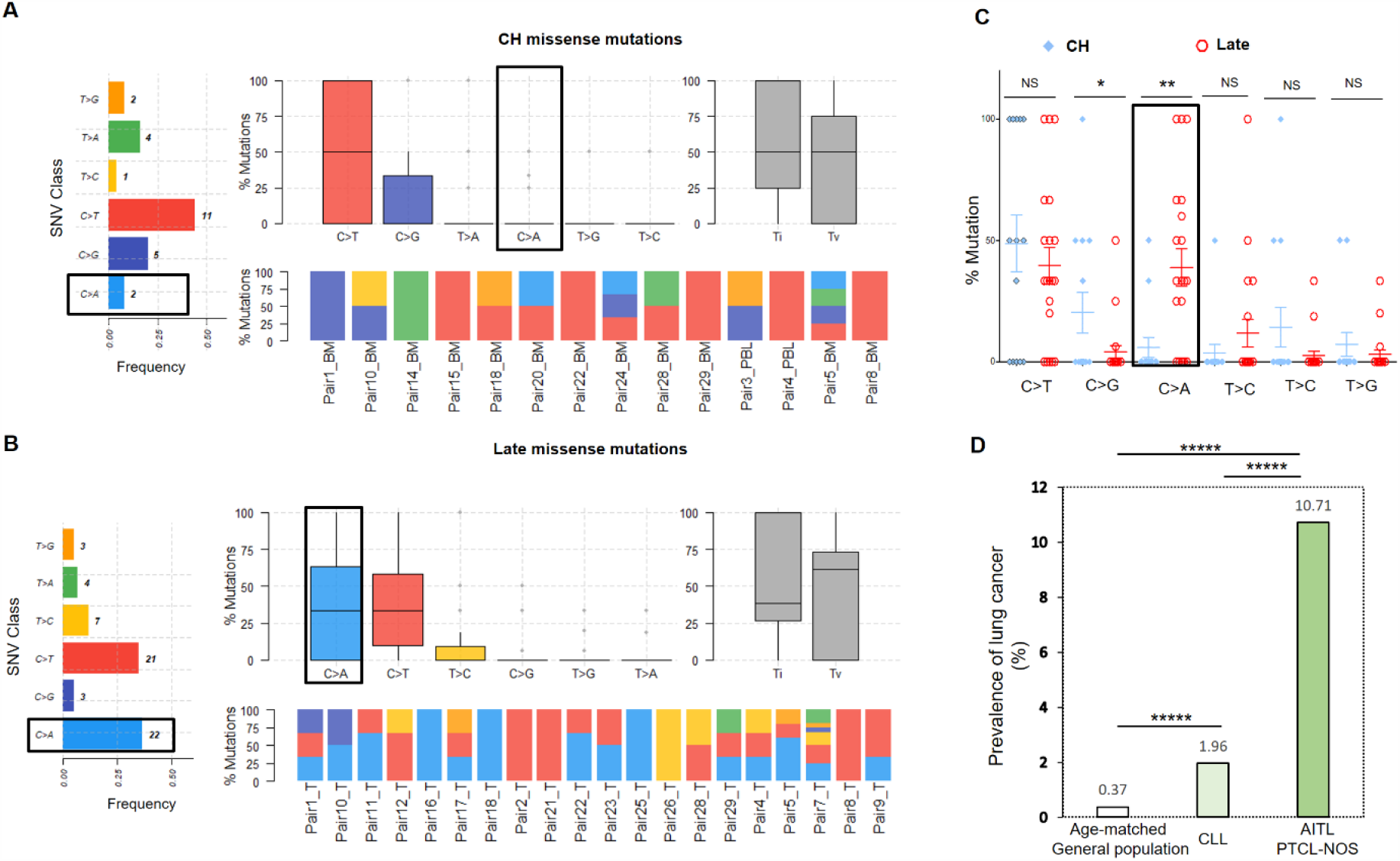
Late mutations in AITL/PTCL, NOS are enriched for C>A transversion substitutions possibly associated with smoking. Titv plot showing overall distribution of the six types of substitutions in the CH (A) and late (B) missense mutations acquired during AITL/PTCL development, as well as fraction of these substitutions in each sample. The median is indicated by a horizontal line. Bar plot on the left showing SNV classes and fraction of each substitution class among all missense mutations. Ti, transitions, Tv, transversions, SNVs, single nucleotide variants. (C) Side by side comparison of transition and transversion base substitutions acquired between the early CH associated and late mutations. (D) Bar plot comparing the prevalence of lung cancer among three populations indicated. * p < 0.05; ** p < 0.01; *** p < 0.001; ***** p < 0.00001; NS, not significant.

In the late mutations, Ti and Tv are also not significantly different (Ti vs Tv, median, 38.53 % vs 61.43 %, mean, 49.31% vs 50.68%, P value >0.05), At the base substitution level, however, besides C>T (35%), C>A emerges as one of the predominant mutant forms (36.7%), which is believed to be associated with the smoking-induced mutational process (**Fig. 2B**) ^23^. On a case basis, C>A substitutions are enriched in late mutations compared to CH mutations (mean, 38.8% vs 5.95%; median, 33.3% vs 0%; t test, P value = 0.0024) (**Fig. 2C**), and the fraction of the cases with C>A substitution in the late mutations is 4-5 times that with C>A in the CH associated mutations (67% vs 16%), suggesting a potential role of the smoking-associated C>A mutational process for acquisition of additional driver mutations in the pathogenesis of AITL.

Enrichment of smoking-associated C>A substitution in the late mutations raises the possibility that patients with AITL/PTCL-NOS might have a higher risk for lung cancer or other smoking associated cancers. To test this hypothesis, we compared the prevalence of lung cancer in three populations, including patients with AITL/PTCL-NOS (the current study cohort, n=28, including one additional PTCL-NOS case without matched PB/BM matched control, Patient #27, see **Supplementary Table1**), patients with chronic lymphocytic leukemia (CLL, n= 1329) and aging-matched general US population (n= 153798075, estimated, age, 40 to 80+ years old) as control group. The data for CLL group was published ^24^, and the data for the control group was downloaded from the CDC website (https://gis.cdc.gov/Cancer/USCS/DataViz.html). Analysis shows that the prevalence of lung cancer in AITL/PTCL-NOS is 27.8 times higher than that in the aged-matched general population (10.71% vs 0.37%, P<0.00001), and 5.47 times higher than that in the CLL group (10.71% vs 1.96%, P<0.00001) (**Fig. 2D**), supporting the association between AITL/PTCL-NOS and smoking-induced tumorigenesis.

## AITL with hematologic neoplasms of other lineages arise from common mutated hematopoietic stem cell precursors

Three patients with AITL presented with additional hematologic neoplasms of other lineages. We present here the clonal evolution patterns of these tumors based on the results of the mutation profiling (**Fig. 3**). One of these cases provides genetic evidence for the progression of CH to overt myeloid malignancy through acquisition of additional mutations (patient #5, **Fig. 3A**). Patient #20 illustrates that the AITL does not necessarily have to be the initially diagnosed malignancy in patients with both AITL and a second malignancy. The third case was an unusual case in which the patient (#14) had CH, AITL as well as DLBCL, the latter was associated with acquisition of an *EZH2* hotspot mutation. Detailed descriptions of these illustrative cases are provided in **Supplementary Text**.

**Fig. 3.**
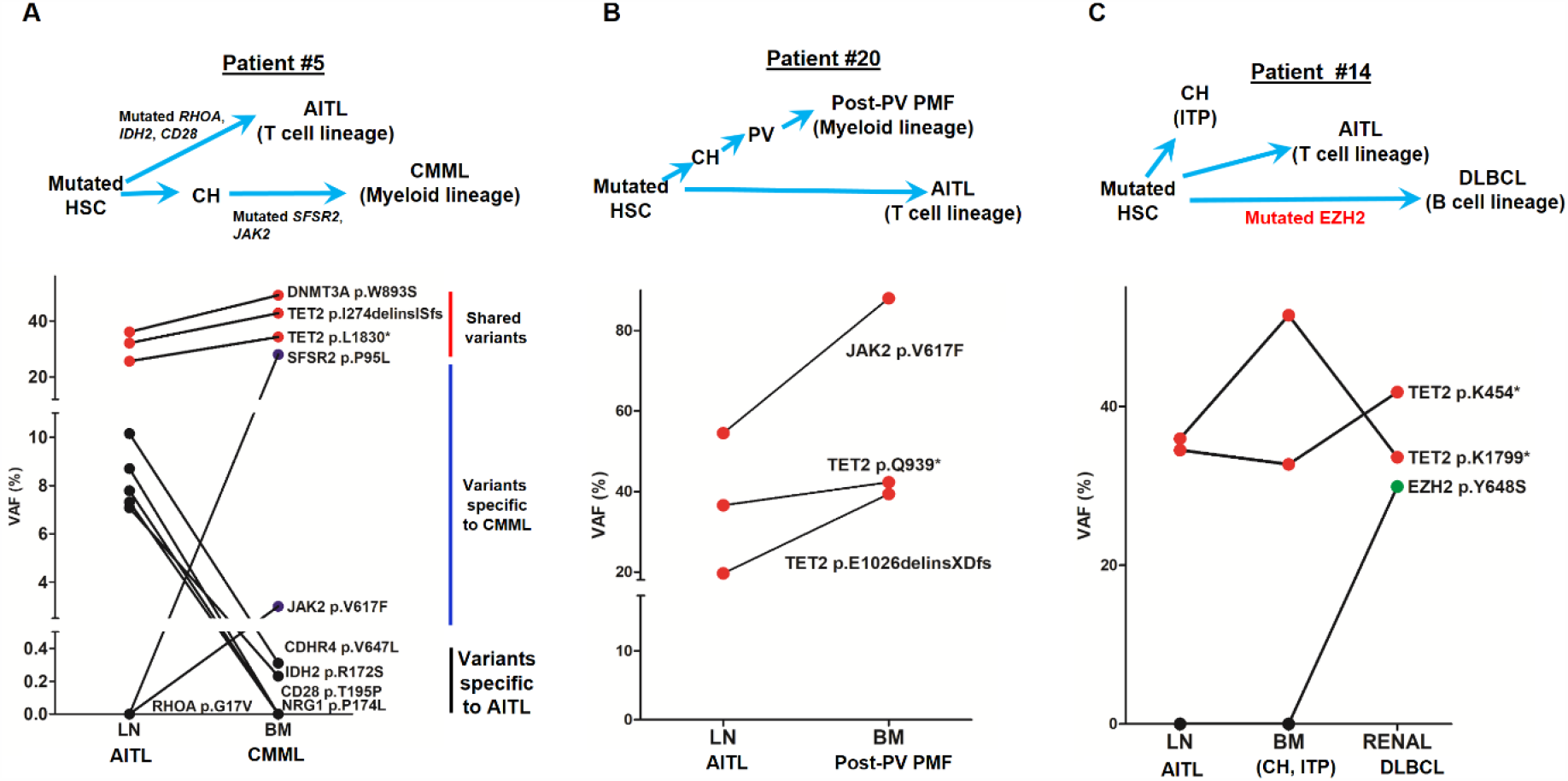
AITL and concomitant hematologic neoplasms develop from common mutated hematopoietic stem cells. (A), (B), (C) Dot plots comparing VAFs of the mutations identified in the AITL and the concomitant hematological malignancies. Red dots show the variants shared between different hematologic neoplasms or entity in the same patient. Dark blue dots in (A) indicate the variants specially related to CMML, and the black dots in (A) denote the AITL-specific variants. Schematic diagrams depicting hypothetical clonal evolution models of the tumors deriving from mutated HSC are also presented. In patient #5 (A), additional mutations besides the CH-associated mutations were identified and implicated in the disease progression to AITL and CMML, respectively. In patient #20, no additional mutations besides those mutated in HSC are identified. In patient #14, a mutated *EZH2*, indicated by green dot, is implicated in the progression to DLBCL. In all 3 cases, there are mutations that are shared between the AITL and the concomitant myeloid or B lymphomas, supporting evolution of these neoplasms from a common precursor. CMML, chronic myelomonocytic leukemia; PV, polycythemia vena; Post-PV PMF, post-PV primary myelofibrosis; DLBCL, diffuse large B cell lymphoma; ITP, immune thrombocytopenia.

Together, our data further provide evidence that AITL can be associated with the development of a hematopoietic neoplasm of different lineages, i.e. myeloid or B-lymphoid, either preceding or subsequent to the diagnosis of AITL. In all cases, truncal mutations common to all lineages are seen, with late mutations seen in specific tumors (e.g. *SRSF2* in myeloid, *EZH2* in DLBCL)

### Impact of destructive *TET2* mutations on development of multiple hematologic malignancies

One of the features shared among the 3 cases with concomitant hematologic neoplasms is that they all had multiple (>1) pathogenic mutations in *TET2*. This observation prompted us to investigate the relationship between *TET2* mutation status and occurrence of multiple hematologic malignancies, specifically through assessing effects of *TET2* mutation status on concomitant hematologic malignancy-free survival in AITL patients. To increase the power of the statistical analysis, the patients included in our study were combined with an outside cohort of AITL patients whose relevant genomic and survival data were recently published ^25^, leading to the total number of 49 cases for survival analysis.

The patients were initially divided into two groups: wild type *TET2* group (no *TET2* mutation found in the BM/PB samples) and pathogenic *TET2* mutant groups (one or more *TET2* mutation detected in the BM/PB samples). Although there was a trend that AITL patients with the pathogenic *TET2* mutations detected in the BM/PB had a worse clinical outcome, no statistically significant differences in the second hematologic malignancy-free survival were observed (P value = 0.3273, stratified hazard ratio = 0.29), consistent with the literature ^26^.

We further stratified the patients into the high *TET2* mutation burden and no or low *TET2* mutation burden subgroups. The criteria for inclusion in the first subgroup (n =8) are as follows: (1) the CH identified in the BM/PB harbored two or more pathogenic mutations in *TET2*, including pathogenic SNVs, nonsense and frameshift mutations interpreted as “Tier 1” or “Tier 2” mutations according to a published professional guideline in molecular pathology ^27^; (2) The VAF of each of the pathogenic *TET2* variants described in (1) was ≥ 15%. The cases that did not meet these two criteria were assigned to the second subgroup (n=41). Kaplan-Meier survival analysis showed that AITL patients carrying two or more pathogenic *TET2* mutations with high allelic burden (the first subgroup) had significantly shorter second hematologic malignancy-free survival (P value = 0.0034) (**Fig. 4**). Cox proportional hazards model also estimated a significantly higher hazard ratio (HR) in the first subgroup (stratified hazard ratio, 10.81, 95% CI of ratio, 2.2 to 53.2). Further analysis shows that specificity and sensitivity of this CHN biomarker reach 97.44%, 70%, respectively, with 87.5% of positive predictive value (PPV) and 92.7% of negative predictive value (NPV). This survival analysis indicates that harboring two or more pathologic mutations in *TET2* with relatively high allele burden (≥15%) is an independent risk factor to predict the second hematologic malignancies in AITL patients.

**Fig. 4.**
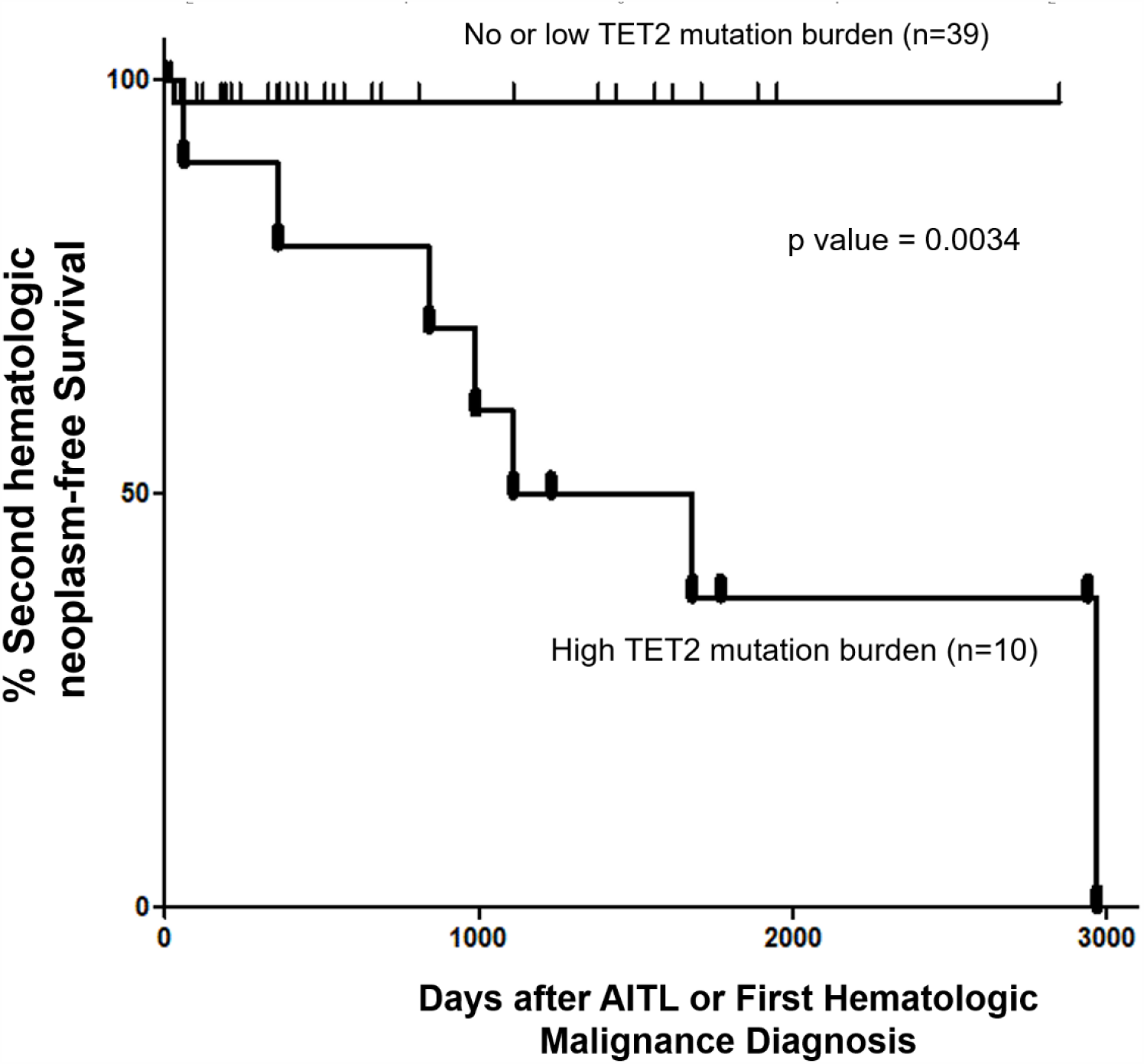
Pathogenic *TET2* mutation status in the BM/PB samples is a predictive biomarker for concomitant hematologic neoplasms in AITL/PTCL-NOS patients. Kaplan-Meier survival curve analysis of concomitant hematologic neoplasm-free survival in AITL or AITL-related patients based on *TET2* mutation status in the BM/PB. Concomitant hematologic neoplasm-free survival of AITL patients can be stratified based on absent/low or high TET2 mutation burden subgroups. p-value was calculated by log-Rank test. In one case, the second hematologic malignancy (PV) preceded the development of AITL.

## Discussion

In the current study, we examined the landscape of the genomic alterations in AITL/PTLC-NOS and their paired BM or PB using a large-panel targeted sequencing approach in the largest cohort of the AITL patients reported to date. We demonstrated that in about 60% of AITL/PTLC-NOS patients, identical pathogenic *TET2* and/or *DNMT3A* mutations were shared between AITL/PTCL-NOS and CH found in the BM or PB. Studies of large cohorts have demonstrated an increased risk of hematologic malignancy for CH ^22,28^, but no definitive link has been established between CH and AITL/PTCL-NOS from those studies. Our findings suggest that these *TET2* and/or *DNMT3A* mutations occur very early in the HSC before they give rise to the common lymphoid progenitors and common myeloid progenitors, and propose a possible link between CH and development of AITL^2^. Interestingly, the VAF of the CH-associated mutations is 22.5% on average in our cohort, and is higher compared to the average VAF of CH-related mutations in the general population ^22^. This observation is in line with the higher risk of hematopoietic malignancy associated with increased VAF (>10%)^29^. It is conceivable that certain *TET2* or *DNMT3A* mutations are stronger drivers which can result in more expanded CH and/or higher efficient T-cell lymphoma development. For example, as seen in patient #28, there were 3 *TET2* mutations identified in the LN, each of which appears to be present in separate clones and have different capacity to generate CH based on VAF in the BM (0%, 5.47% and 10.89%, respectively). In addition, our study supports a mutated HSC origin for AITL. As the *TET2* and/or *DNMT3A* mutations are propagated to the lymphoid and myeloid progeny of the mutated HSC, it can be speculated that in the lymphoid compartment, the impacts of these mutations vary depending on the developmental and differentiation stage of the T-cells, and may be most felt in the T-cells of follicular helper cell origin (TFH). Lastly, our interesting case of an AITL patient with CH and subsequent development of DLBCL and the sharing of the same TET2 mutation among all three lesions suggest that a subset of DLBCL, possibly the molecular subtype characterized by mutated *TET2* ^30^ may originate from mutated HSC. To our knowledge, this is the first reported case in which the mutated HSC developed into three distinct tumors of diverse lineages.

The findings from this investigation confirm and extend the results previously published regarding the cellular origin of AITL ^12,21,25,31-34^. Most of these previous studies presented sporadic AITL cases in which the *TET2* or/and *DNMT3A* mutations present in AITL were also found in their BM/PB compartments. Two reports showed that AITL shared the same *TET2* mutations with the isolated CD20^+^/CD19^+^ (B cells) or CD34^+^ cells ^31,33^. These studies also pointed to a mutated HSC that gives rise to lymphoid and myeloid cells harboring the same mutations. A high-risk CH was also documented as the cellular origin of AITL and *NPM1*-mutated AML in a patient ^21^. While our manuscript was under preparation, the results of a study conceptually similar to ours regarding the cellular origin of AITL was reported ^25^. Consistent with our observations, the report showed that the mutations related to CH (i.e. *TET2* or *DNMT3A*) were detected in both the neoplastic T-cell and myeloid compartments in 15 out of 22 AITL patients (68%), and associated with second myeloid neoplasm development after the diagnosis of AITL in 4 cases. However, in their cohort, no cases were reported where AITL developed subsequent to myeloid neoplasms. Our study presented one such case (Patient #20) whose AITL developed after 10 years of PV and the two hematologic neoplasms shared three identical *JAK2* and *TET2* mutations (**Fig. 3B**). The identification of cases in which myeloid neoplasms precede the diagnosis of AITL provides additional supportive evidence to the postulation that the mutated HSCs are the common origin for these hematologic neoplasms, which develop independently and divergently in tumor evolution. Whether ATIL precedes or develops subsequent to the myeloid neoplasms may depend on the stochastic dynamics of the clonal evolution.

Additional late non-CH mutations are found in 68.4% of the AITL/PTCL-NOS in our cohort, consistent with the belief that CH-associated *TET2* and *DNMT3A* mutations are insufficient for tumorigenesis and additional genetic alterations are required. Consistent with this notion, in AITL animal models, *TET2* disruption or *RHOA*^*G17V*^ expression alone failed to induce AITL development; however, AITL-like lymphoma developed once *TET2* disruption and *RHOA*^*G17V*^ expression were combined^35-37^. For the development of myeloid neoplasms, additional mutations beyond CH-associated *TET2* and *DNMT3A* mutations drive further clonal expansion from CH. These mutations may be acquired early (patient #20, *JAK2*, Fig, 3B) or late during tumor development (patient # 24, *JAK2*, **Supplementary Fig. 2**).

We discovered that the late non-CH mutations are enriched for the missense mutations and the C>A substitutions (**Fig. 1E, Fig. 2**). In the current cohort, the C>A base substitution is associated with critical mutations in a number of oncogenic genes. This finding may have implications on treatment and prevention of AITL. It is believed that the C>A mutation is likely caused by mis-replication or mis-repairing of DNA damage induced by tobacco carcinogens ^23,38^, which largely result in missense mutations^39^. Consistent with this causative link is our findings that the majority of the late non-CH mutations identified in our cohort were missense mutation (75.2%, **Fig.1E**), and that the patients with AITL/PTCL-NOS have a 27.8-fold increased risk for development of lung cancer compared to the age-matched general population (**Fig. 2D**). Consequently, our findings suggests that cessation of smoking may be a potential effective intervention to prevent AILT development in higher risk population, particularly those already found to harbor CH.

On the contrary, the CH-associated genetic alterations in the current cohort are characterized primarily by the C>T and C>G mutations (64% of all the mutations) in *TET2* and *DNMT3A* (**Fig.2A, 2C**). This pattern was reported to be associated with the AID/APOBEC family of cytidine deaminases or aging-dependent function decline of base-excision repair machinery ^23^. This mutational mechanism might also play a role in the non-CH late mutations, as 35% of the non-CH mutations were C>T substitutions (**Fig. 2B, 2C**). Interestingly, reduced accumulation of the C to T mutations by inactivation of *AID* blocked development of B cell malignancies in aging *TET2* deficient mice^40^, implying that AID might be a therapeutic targeting candidate for lymphoma, including AITL.

Furthermore, we found that CH associated with multiple-hit *TET2* (defined as ≥2 pathogenic *TET2* mutations with VAFs of ≥ 15%) is an independent risk factor for development of concurrent hematologic malignancies (Fig. 4). Recently, certain features of CH predictive of hematopoietic malignancy development were identified^29^. These features include >1 mutated gene, VAF≥ 10% and mutations in specific genes and variants, for example *TP53* and *IDH1/2*Our *TET2* biomarker includes 2 or more mutations and VAF of at least 15%. However, *TET2* has not been previously implicated as a marker for increased risk of hematologic malignancy in CH in general. It is possible that this multiple-hit TET2 biomarker is specific and only relevant in the setting of patients with AITL and CH. Mechanistically, it is possible that two or more *TET2* mutations each with relatively high mutation burden (≥15%) correlates with increased clonal expansion and/or more severe disruption of TET2 activity, thereby increasing the global chance of acquiring additional driver mutations and hence increased risk for development of second hematologic neoplasms. Consistent with this hypothesis, aging *TET2* deficient mice develop diverse hematologic malignancies ^41^. A reliable predictor for concurrent hematologic malignancies may be helpful for clinical stratification and management for this subset of the AITL patients. For myeloid malignancies with the double-hit *TET2* mutations, more intensive therapeutic regimen like bone marrow transplantation might be warranted for reducing the risk of AITL development.

In summary, our study provides genomic evidence of an origin of AITL/PTCL-NOS from a mutated HSC clone, which can be associated with CH as well as development of myeloid and even B-cell malignancies. The development of these hematopoietic malignancies of different lineages occur via divergent evolution from the mutated HSC clone, often with acquisition of additional mutations frequently induced by mutagenic agents like tobacco. We also identified a potential biomarker: two or more pathogenic TET2 mutations with high mutation burden for the development of second hematologic neoplasm in AITL patients. Single cell methodology will help definitively determine the clonal architecture in lymphoma and BM with multiple mutations and enhance our understanding of the initiation of tumor induced by CH-associated mutations.

## Materials and Methods

### Patients and Study Samples

All tissue samples (27 lymph node tissue specimens, 27 bone marrow aspirate/peripheral blood samples) were collected from 25 AITL or 2 PTCL NOS patients who were diagnosed or confirmed from June 2010 to December 2019 following World Health Organization classification criteria by attending hematopathologists at NYP/Weill Cornell Medical Center, and clinical Information was obtained from electronic clinical records. Of these 27 study cases, 4 were initially diagnosed with PTCL with THF phenotype (**Supplementary Table1**), and included as the part of the AITL cases according to their similar clinical and molecular features as recently proposed by WHO^8^. This study was conducted in accordance with the Declaration of Helsinki regulations of the protocols approved by the Institutional Review Board of Weill Cornell Medicine, New York, USA. Written consent for use of the samples for research was obtained from patients or their guardians.

Genomic DNA was extracted from lymph node tissue and bone marrow or PBMC cell pellets following manufacturer’s instructions (QIAamp DNA Mini Kit, Qiagen, Germantown). DNA samples and sequencing libraries used in targeting sequencing as described below were quantitated by Tape Station (Agilent Technologies, Santa Clara) and Qubit (Thermo Fisher Scientific, Singapore).

### T Cell Targeted Sequencing

A 538 gene targeted sequencing panel were designed to investigate the genomic profile of the primary tumors and the BM/PB tissues^20^. The genomic regions covered by sequencing include coding exons and splice sites of these genes (target region: ∼3.2 Mb) that were reported being recurrently mutated (>2) in mature T-cell neoplasms, as well as genomic regions corresponding to recurrent translocations. Using an input of genomic DNA of at least 100 ng isolated from frozen tissues or FFPE samples, the next-generation sequencing (NGS) libraries were constructed using the KAPA Hyperplus Kit (Roche, Basel, Switzerland), and hybrid selection was performed with the Twist Library Prep Kit (Twist Biosciences, San Francisco, CA, USA), according to the manufacturer’s protocols. Multiplexed libraries were sequenced using 150-bp paired end Hiseq4000 sequencers (Illumina, San Diego, CA, USA). NEXTGENe software (Softgenetics, State College, PA, USA) was used to perform bioinformatic analysis (SNV and INDEL variant calls) with standard settings recommended by the manufacturer. Specifically, cutoff values for the variant allele frequency, population frequencies, and strand bias were set at 5%, 0.01%, and 1:5, respectively.

### Myeloid NGS Panel

Targeted enrichment of 45 genes recurrently mutated in myeloid malignancies was performed using the Thunderstorm system with a customized primer panel^42^. The primers target coding exons of the genes, leading to a total of 726 amplicons. Libraries were prepared by microdroplet-based PCR target enrichment method from DNA, followed by sequencing using the Illumina MiSeq yielding 260-bp paired end reads. Sequencing data were analyzed and reported with a customized analytical pipeline. This NGS panel testing is performed in a clinical lab CLIA-certified and accredited by the College of American Pathologists.

## Data Analysis

Data analysis were conducted with GraphPad/Prism 5 software and various R packages, including base packages, ggplot2, ComplexHeatmap and Maftools. The survival comparison was analyzed using Kaplan-Meier curves (log-rank test, significance defined as p < 0.05).

## Data Availability

All data referred to in the manuscript are provided or available upon request.

## Abbreviations

AITL: angioimmunoblastic T cell lymphoma
PTCL-NOS: peripheral T cell lymphoma, not otherwise specified
CH: clonal hematopoiesis
CHN: concomitant hematologic neoplasm
DLBCL: diffuse large B cell lymphoma
MPN: myeloproliferative neoplasm
TET2: Ten-Eleven Translocation-2
VAF: variant allele frequency
Ti: nucleotide transition substitution
Tv: nucleotide transverse substitution
PPV: positive predictive value
NPV: negative predictive value

## Funding

This study is supported by a grant (R01 CA194547) from the National Cancer Institute to WT.

## Competing interests

The authors declare no competing financial interests.

## Contributions

SC contributed to the study design and conceptualization of the project, optimized bioinformatics pipeline, analyzed and interpreted the data, and wrote the manuscript. WZ performed the experiment and contributed to the data analysis. GI provided critical study materials and contributed to the revision of the manuscript. WT conceptualized and directed the project, designed the experiments, analyzed and interpreted the data, and wrote the manuscript.

## Supplementary Materials

## Supplementary Text

**Fig.1A: 6 illustrative cases in which common mutated HSC developed into CH and AITL or PTCL-NOS**

In Patient #1, identical *TET2* and *DNMT3A* somatic mutations, p. E1089fs and p.F791L, were identified both in CH (VAFs: 48.39%, 34.62%) and in the neoplastic T cells in the diagnostic LN specimen (VAFs: 36.87%, 27.59%) (**Figure 1A**). The other four pathogenic variants, *TET2* p.Q743*, *RHOA* p.G17V, *IDH2* p.R172T and *PTPRF* p.R654H, were detected at high VAFs in the LN but at much lower VAFs in the BM (**Figure 1A**). These results demonstrate that the latter four mutations are likely acquired at a later time point during AITL development (late mutations) and the low-VAF variants detected in the BM represent minimal involvement in the BM and not CH. Patient #4 had no overt evidence of a myeloid neoplasm in the BM while diagnosed with AITL, and had 2.1% bone marrow involvement by AITL according to immunophenotypic and gene rearrangement studies. The T cell NGS panel identified 7 pathogenic mutations in the primary lymphoma, of which 6 were also found in the matched BM. The allelic burden of the two mutations with the highest VAFs (*DNMT3A* p. Q678*, VAF=6.22%; *ATP1A3* p.V216M, VAF=4.47%), were 2 to 3 times that of the estimated tumor burden (2.1%) in the BM, suggesting that *DNMT3A* p. Q678* and *ATP1A3* p.V216M were not only present in the neoplastic T-cells but also associated with CH. The other 5 mutations, including *RHOA* p.G17V and *IDH2* p. R172G, were found at the VAFs ranging from 0% to 1.5% in the BM, consistent with lymphoma involvement rather than CH. These results suggest that these 5 mutations were acquired subsequent to the CH-associated mutations described above. Interestingly, sequencing a relapsed lymphoma specimen from this case identified only one mutation *DNMT3A* p. Q678*. This mutation information in the relapsed lymphoma helps clarify the clonal architecture of both the primary AITL and CH. First, it suggests that the primary AITL harbored two clones, a dominant clone with *ATP1A3, DLGAP3, TET2, RHOA* and *IDH2* mutations, and a minor clone with *DNMT3A* mutation, the latter being the precursor clone for relapse. Second, it also implies that there are 2 clones in CH, one associated with the *DNMT3A* mutation and the other with the *ATP1A3* mutation. In Patient #10, the CH-associated variant shared between the primary lymphoma and the matched BM, *TET2* p. N1484K, was identified at high allelic burden in both the lymphoma and BM samples (VAFs in LN vs BM, 47.33% vs 38.26%,). Another *TET2* mutation, p. D1378G, was also identified in the both the LN and the BM, at VAFs of 16.89% and 0.32% respectively. The large difference of the VAF in the BM of the two *TET2* mutations suggests that they may belong to different HSC clones, each of which has markedly different CH contribution, or the *TET2* p.D1378G mutation was acquired subsequent to the *TET2* p.N1484K mutation as a subclonal mutation in the HSC. The other 3 mutations: *TET2* p.V415fs, *RHOA* p.G17V and *IDH2* p.R172T were present exclusively in the lymphoma sample. In patient #29, the *DNMT3A* R882H hot-spot mutation was shared at high VAFs in both the lymphoma and BM (46% and 35.6%), consistent with a CH-associated mutation, while the other 4 mutations detected in the LN was present in the BM at low VAFs, consistent with BM involvement by T-cell lymphoma. Interestingly, both the *TET2* N1484K mutation in patient #10 and the *DNMT3A* R882H mutation in patient #29 were present in the primary lymphomas at a VAF of close to 50%, about 3-4 times the allelic fractions of other mutations identified in the LN. This finding implies the presence of these mutations in almost the entire cell population in the LN. However, based on immuno-morphologic evaluation and the VAF of the other mutations identified in the LN, the tumor burden in the LN for these two cases is about 20-40%. This raises the possibility that besides the neoplastic T cells, reactive lymphocytes in these two cases might also harbor the CH-associated mutations.

In the two PTCL-NOS cases (#2 and #18), we observed similar findings as described above for AITL (**Fig. 1A**). Patient #2 showed eosinophilia with 1.5% neoplastic T cells involvement in the BM. Two pathogenic *STAT3* mutations were identified. One could be considered a CH-associated mutation (*STAT3* p.Y657_K658insALL, VAFs in LN vs BM, 21.61% vs 8.34%) since its VAF is considerably higher than the estimated percentage of tumor involvement in the BM. The other is most likely PTCL-NOS-related mutation (*STAT3* p.W474*). This *STAT3* nonsense mutation had the allelic burden of 18.33% in the primary lymphoma but was present at much lower level (VAF = 2%) in the matched PB, in line with the estimated tumor burden. Patient #18 presented mildly hypercellular marrow with mild granulocytic hyperplasia while diagnosed with PTCL-NOS and had no involvement in the BM per the overall pathologic studies. The NGS target panel identified a splice mutation in *TET2* (c.3501-1G>A) and a missense mutation in *SETX* (p. Y2258D) shared between CH and PTCL-NOS. Two mutations (*TP53* p.P151T and *ARID1A* p. G779*) were only found in the neoplastic T cells, presumably representing later mutations in PTCL-NOS development following acquisition of the *TET2* and *SETX* mutations.

**Fig. 3: three AITL cases with concurrent hematologic neoplasm**

Patient #5 was initially diagnosed as AITL, followed by a diagnosis of chronic myelomonocytic leukemia (CMML) 7 months later. In the BM specimen, taken four months prior to the diagnosis of CMML and sequenced in this study, the overall pathologic findings showed 1-5% involvement by AITL and suspected involvement by a myeloid neoplasm. Three identical mutations, including 2 *TET2* and one *DNMT3A* mutations: p.I274delinsISfs, p.L1830* and p.W893S alterations, respectively, were identified in both the LN and BM with high allele burdens (VAF range, 25-50%, **Supplementary Table2**). In addition, pathogenic *SRSF2* and *JAK2* mutations were identified in the BM. These findings support a scenario in which AITL and CMML arose via divergent evolution from a common HSC clone mutated in the CH-associated *TET2* and *DNMT3A* genes. Subsequent development involves accumulation of additional mutations: *SRSF2* and *JAK2* mutations from CH to CMML, and *IDH2* with other mutations to AITL.

Patient #20 was initially diagnosed with PV and progressed to post-PV PMF after 10 years. Two months later, the patient was also diagnosed with AITL. We sequenced the T-cell lymphoma in the LN and the paired bone marrow specimen with post-PV PMF (the initial PV specimen was unavailable). The overall pathologic studies showed no lymphoma involvement in the BM. The NGS sequencing revealed that the T and myeloid malignant cells harbored two identical destructive *TET2* mutations with high mutant allele burdens, p. Q939* (20% VAF) and p. E1026delinsXDfs (42% VAF). The driver mutation for MPN, *JAK2* p.V617F, was not only present in the post-PV PMF (88% VAF), but also found in the concomitant AITL with the mutant allele burden of 54.5% (**Fig. 3B, Supplementary Table2)**. No *IDH2* or *RHOA* mutation was detected in the AITL. These findings suggest that the *JAK2* driver mutation was acquired early in the HSC. Indeed, *JAK2* mutation was thought to be an early event in PV [1]. In this particular case, the combination of *TET2* and *JAK2* V617F may also be sufficient to drive AITL development, as no other drivers like *IDH2* or *RHOA* were found.

Patient #14 initially presented as immune thrombocytopenia (ITP) and had a splenectomy in 2008. He was diagnosed with AITL two years later and DLBCL about 7 years later. We sequenced two diagnostic tissue samples for the AITL and DLBCL, respectively, as well as one BM sample collected from this patient in 2010. The morphologic, immunophenotyping and molecular findings confirmed that the BM sample was not involved by AITL or a myeloid neoplasm. Interestingly, two nonsense *TET2* mutations, p.K454* and p.K1799*, were identified in all three samples with the VAFs ranging from 32% to 51%. An additional pathologic mutation *EZH2*, p. Y646S, was detected only in the DLBCL (**Fig. 3C, Supplementary Table2)**. The results from this patient suggest that the mutated HSC clone, harboring *TET2* p.K454* and p.K1799* alterations, differentiates into progeny cells of three different lineages (myeloid, T lymphoid, B lymphoid), each of which gives rise to CH, or transformed to AITL or DLBCL. The additional *EZH2* mutation has been reported in B-cell lymphomas[2, 3] and likely plays a role in the DLBCL development.

---

**Supplementary Fig. 1.**
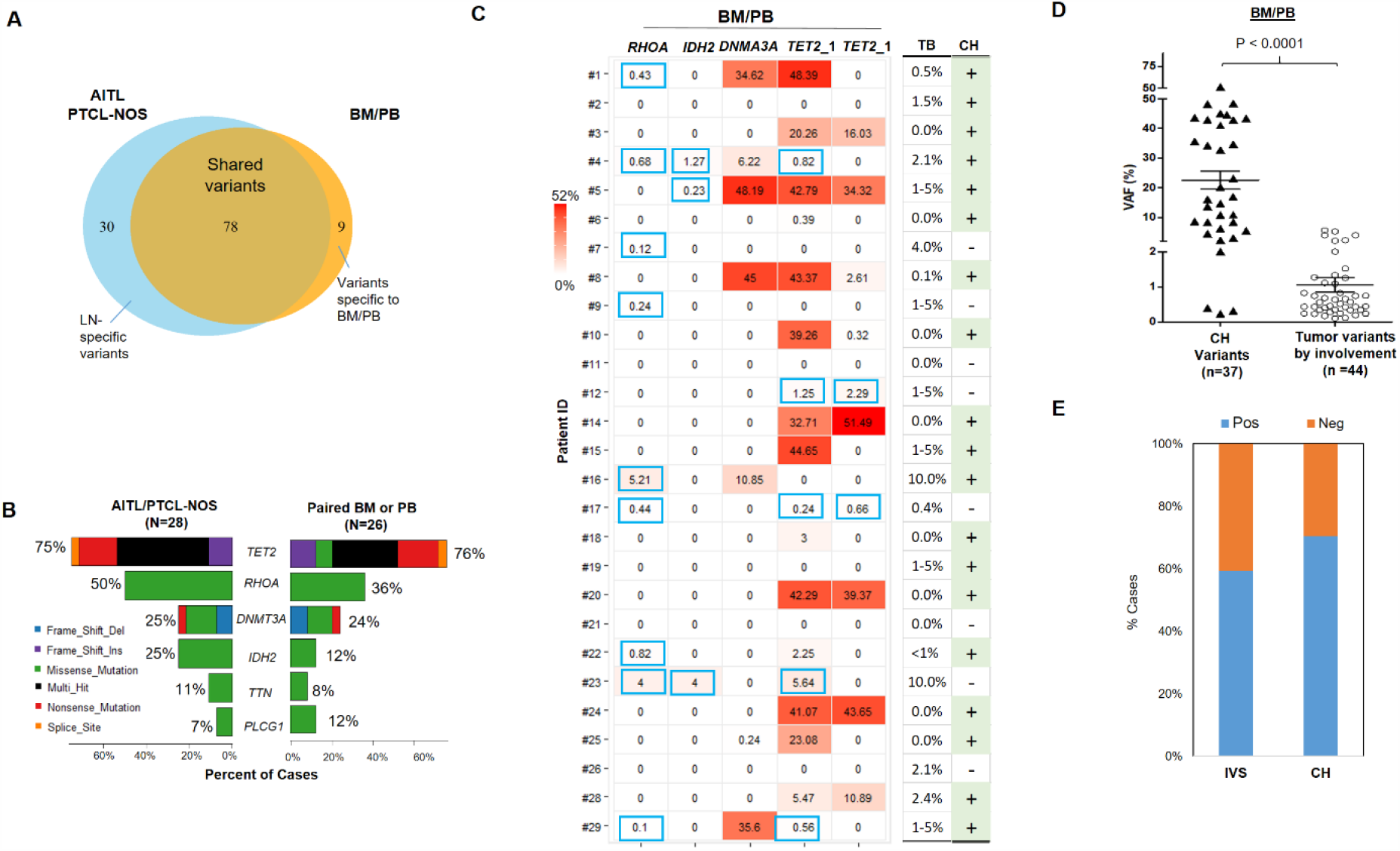
AITL variants involving BM/PB. (A) Venn diagram illustrates the distribution of the shared, and the LN or BM/PB-specific mutations. Note that the variants detected in the BM/PB due to lymphoma involvement are included in this diagram, in contrast to Fig. 1B presented in the main text. (B) The type of variants and mutation frequencies (relative to the total number of cases) for the top 6 mutated genes identified in AITL/PTCL-NOS and matched BM/PB are shown. Different colors represent variant classifications as indicated. (C) Heat map showing the mutations detected in the BM/PB samples in each case. Only the major recurrent mutations are listed. The tumor burden (TB) in the BM/PB was estimated based on morphologic and immunophenotypic and molecular assessment, and the predicted CH status (determined based on comparison of the VAF of the mutations with TB) is indicated. Blue rectangles highlight the VAFs of the variants due to AITL involvement. CH, clonal hematopoiesis; +, presence of CH; -, absence of CH; $, mutation in *STAT3*; ^, mutation in *DDX11*. (D) Dot plot comparing VAFs of the CH-associated variants and those related to lymphoma involvement in the BM/PB specimens. Mean ±SEM of the VAFs is shown for each subgroup. (E) The proportion of the cases with or without BM or PB involvement by the neoplastic T cells (IVS), or with/without clonal hematopoiesis (CH), respectively, is summarized.

**Supplementary Fig. 2.**
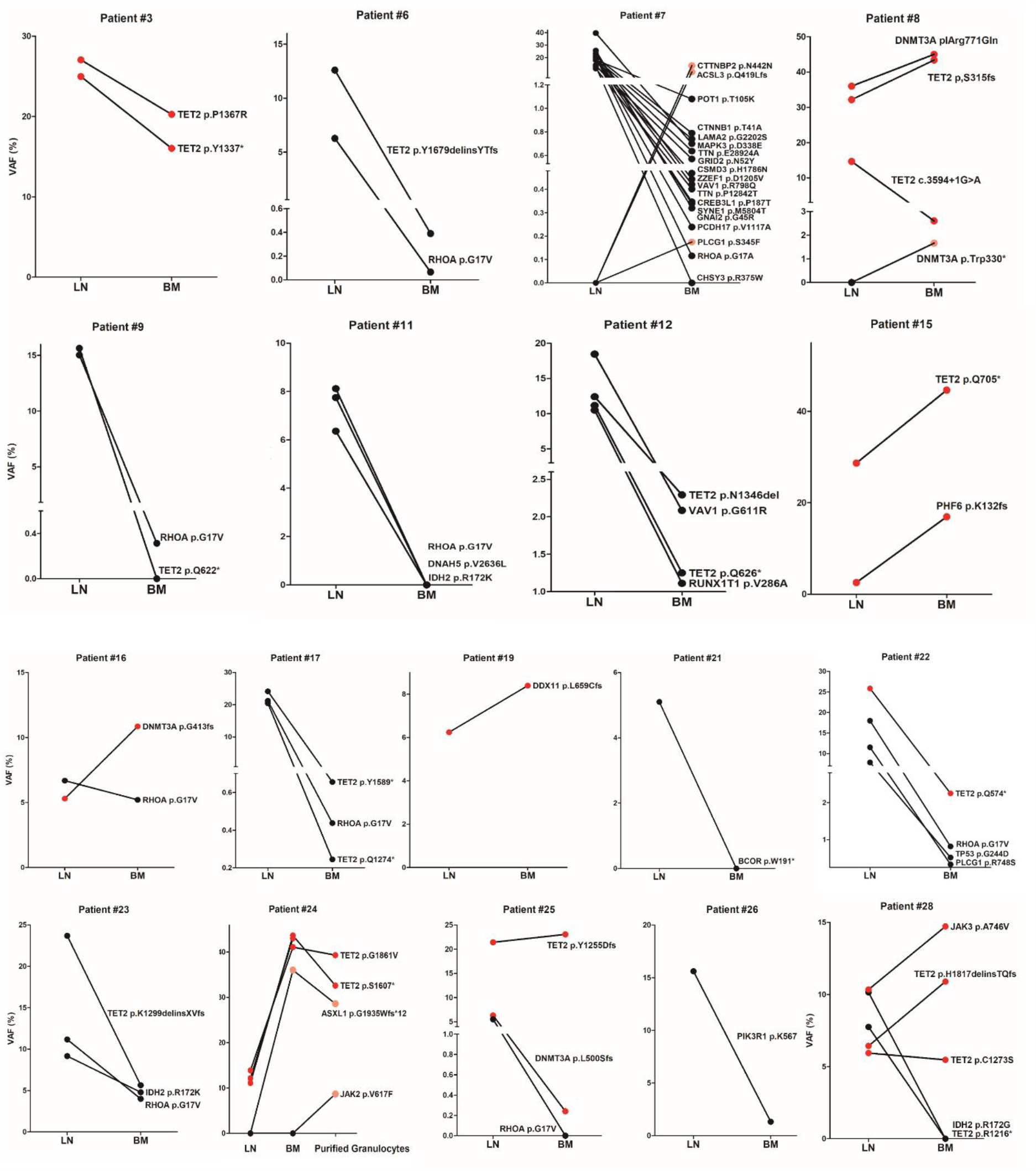
Comparison of VAFs of the variants found in paired AITL and BM/PB samples. Red color highlights the CH-associated variants shared between the LN and BM/PB compartments. The black circles indicate variants specific to the lymphomas, the pink circles represent mutations specific to BM/PB. Cases that were not presented in Fig. 1 and Fig. 3 are shown in this figure. In Pt #7, three mutations specific to BM were found in PLCG1, ACSL3 and CTTNBP2. These may represent novel CH-associated mutations. In Pt #8, DNMT3A p.W330*, likely representing a minor CH clone, was also found specifically in the BM. In Pt.#24, an additional ASXL1 frameshift mutation was identified in the BM, which showed hypercellularity with increased granulopoiesis. In this patient, we appeared to capture the acquisition of a subclonal *JAK2* V617F mutation during tumor progression, which was identified by the myeloid panel in the purified granulocytes from the PB but not detected in the BM one year before (#24).

**Supplementary Fig. 3.**
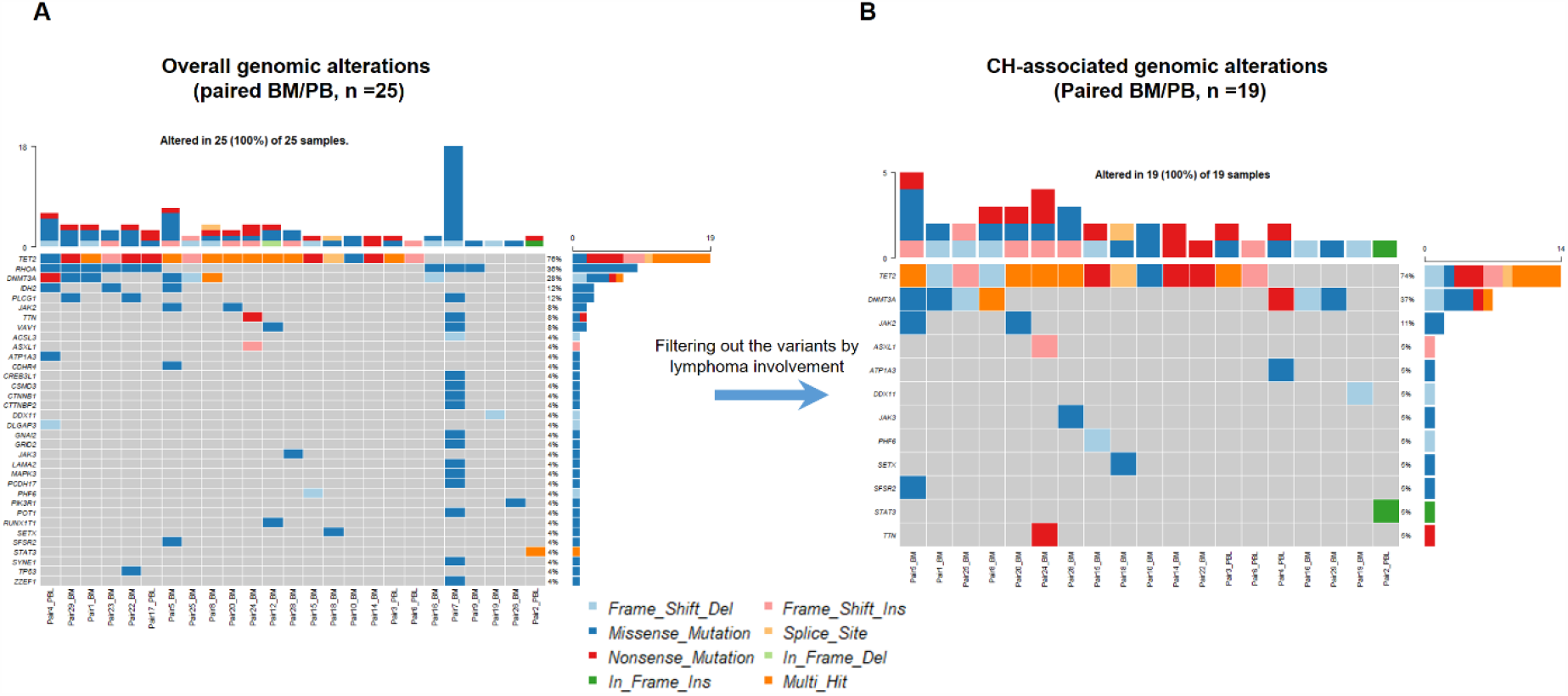
Overall and CH-related genomic alterations in the matched BM/PB samples. (A) The overall mutation profile in the BM/PB shown. (B) The CH–related mutation profile identified in the BM/PB is shown after excluding the variants due to AITL involvement. Top bars on each plot indicate the numbers of the variants detected per sample. The percentage showing gene mutation frequency in the patient cohort is shown at the right. Sample IDs are indicated at the bottom of each plot. Variant classifications are indicated with different colors.

**Supplementary Fig. 4.**
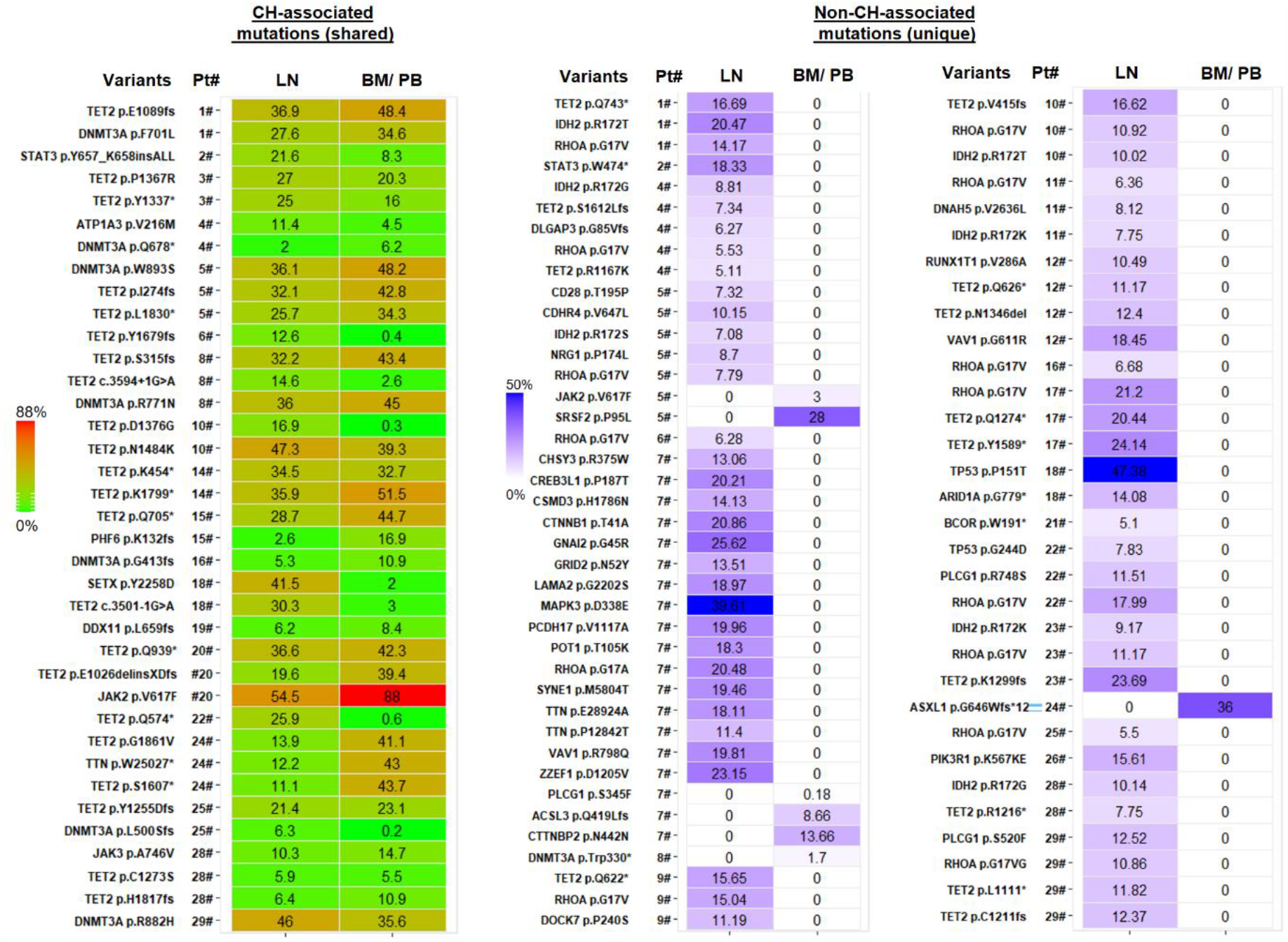
Summary of all CH-associated aned non-CH-associated mutations identified in AITL/PTCL, NOS and their matched BM/PB. **Pt#**, Patient number. Values on the heat maps represent VAFs of the individual mutations.

**Supplementary Fig. 5.**
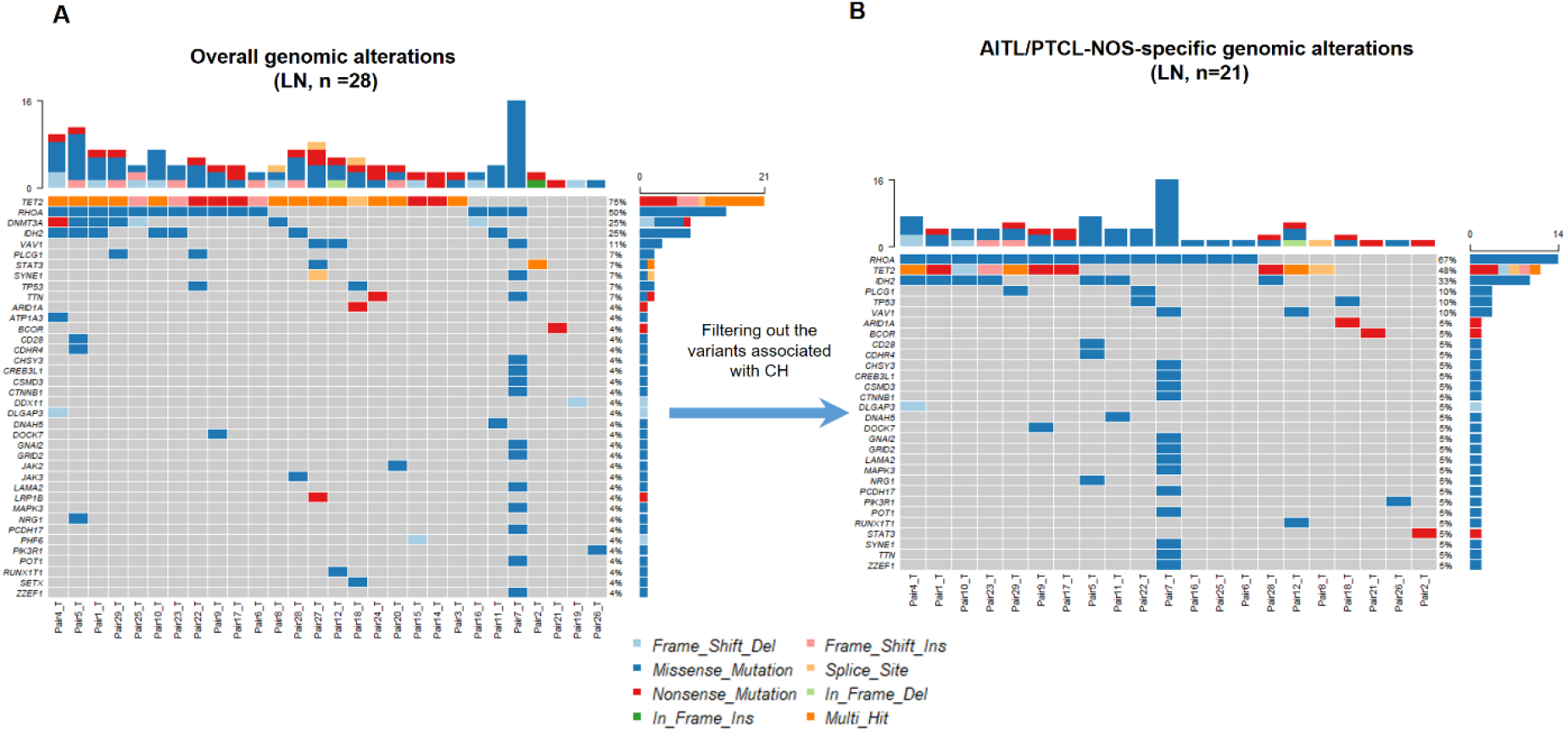
Overall and lymphoma-specific genomic alterations in primary AITL/PTCL-NOS. (A) Mutation plot showing the overall mutation profile in the LN tissues involved by AITL./PTCL-NOS. (B) Mutation plot showing the AITL/PTCL-NOS-specific mutations identified in lymphoma samples after excluding the CH-associated variants. Top bars on each plot indicate the numbers of variants detected per sample. The percentage at right shows the gene mutation frequency in the cohort of the patients. Sample IDs are indicated at the bottom of each plot. Variant classifications are indicated with different colors.

**Supplementary Table 1.** Patient characteristic.

| Patient Number | T | Sex | Age | Dx | Pathology of BM/PBL | Concurrent hematologic Malignancy | Morphology | MFC | TCRG Rearrangement | TB |
| --- | --- | --- | --- | --- | --- | --- | --- | --- | --- | --- |
| #1 | BM | F | 50-59 | Lung cancer, AITL | megakaryocytes slightly increased in number without significant cytologic atypia. PBL, Normocytic anemia. Leukocytosis with neutrophilia | No | + | 0.5% | n/a | 0.5% |
| #2 | PBL | M | 30-39 | PTCL, NOS | BM, hypocellular bone marrow with relative eosinophilia, PBL, eosinophilia | No | - | 1.50% | PCR+ | 1.5% |
| #3 | PBL | M | 50-59 | AITL | unremarkable | No | - | 0 | PCR- | 0.0% |
| #4 | BM | F | 60-69 | AITL | involvement, megakaryocytes are focally clustered and some have atypical nuclear lobation, PBL, unremarkable | No | + | 2.10% | PCR+ | 2.1% |
| #5 | BM | F | 60-69 | AITL | granulopoiesis, dysgranulopoiesis, decreased erythropoiesis, monocytosis, unremarkable T cells. PBL, Absolute monocytosis. | CMML, 10 months post-AITL Dx | - | 0 | PCR+ | 1-5%<sup>\$</sup> |
| #6 | PBL | M | 70-79 | AITL | Megakaryocytes were dysplastic (>10%). Including small / giant forms with hypo / hyperlobulated nuclei. Myelopoiesis is hyperplastic and slightly left-shifted. Mild dysgranulopoiesis is present (<10%) with giant forms and abnormal segmentation. | No | - | 0 | n/a | 0.0% |
| #7 | BM | M | 70-79 | AITL | involvement | No | + | 4% | n/a | 4.0% |
| #8 | BM | M | 70-79 | Prostate cancer, PTCL-TFH | involvement | No | + | 0.10% | n/a | 0.1% |
| #9 | BM | F | 50-59 | AITL | unremarkable | No | - | 0 | PCR+ | 1-5%<sup>\$</sup> |
| #10 | BM | M | 60-69 | AITL | unremarkable | No | - | 0 | PCR- | 0.0% |
| #11 | BM | F | 70-79 | AITL | ITP | No | - | 0 | PCR- | 0.0% |
| #12 | PBL | M | 70-79 | AITL | unremarkable | No | - | 0 | PCR+ | 1-5% |
| #14 | BM | M | 70-79 | PTCL-TFH, DLBCL | ITP | DLBCL, 7 years post-AITL Dx | - | 0 | PCR- | 0.0% |
| #15 | BM | M | 70-79 | AITL | unremarkable | No | - | 0 | PCR+ | 1-5%<sup>\$</sup> |
| #16 | BM | F | 80-89 | AITL | involvement | No | + | 10% | PCR+ | 10.0% |
| #17 | PBL | M | 80-89 | AITL |  | No | - | 0.40% | PCR+ | 0.4% |
| #18 | BM | M | 60-69 | PTCL, NOS | Mildly hypercellular marrow with mild granulocytic hyperplasia | No | - | 0 | PCR- | 0.0% |
| #19 | BM | F | 50-59 | AITL | unremarkable | No | - | 0 | PCR+ | 1-5%<sup>\$</sup> |
| #20 | BM | M | 40-49 | Lung Cancer, post-PV MF, PTCL-TFH | post-PV myelofibrosis | PV, post-PV myelofibrosis prior to AITL Dx | - | 0 | PCR- | 0.0% |
| #21 | BM | F | 40-49 | AITL | unremarkable | No | - | 0 | PCR- | 0.0% |
| #22 | BM | F | 60-69 | AITL | a mildly hypercellular bone marrow with myeloid hyperplasia | No | - | 0 | PCR- | <1% |
| #23 | BM | M | 60-69 | DLBCL and AITL | unremarkable | DLBCL | + | 10% | PCR+ | 10.0% |
| #24 | BM | M | 30-39 | AITL | Myelopoiesis is hyperplastic and slightly left-shifted with ~4% blasts. Evidence of significant dysplasia is not seen | No | - | 0 | PCR- | 0.0% |
| #25 | BM | M | 70-79 | AITL | Hypercellular bone marrow with myeloid hyperplasia | No | - | 0 | PCR- | 0.0% |
| #26 | BM | M | 60-69 | PTCL-TFH | involvement | No | + | 2.10% | PCR+ | 2.1% |
| #27 | no matched BM or PBL | F | 60-69 | Lung cancer, PTCL, NOS | n/a | n/a | n/a | n/a | n/a | n/a |
| #28 | BM | F | 70-79 | AITL | involvement | No | + | 2.40% | PCR+ | 2.4% |
| #29 | BM | M | 50-59 | CTCL to AITL | small population of abnormal myeloid blasts noted in BM, but no MPN diagnosed | No | - | 0 | PCR+ | 1-5%<sup>\$</sup> |

T, type of specimen TB, neoplastic T cell burden by BM involvement CH, clonal hematopoisis \$ limit of detection of TCRG (1-5%) n/a, data unavailable DLBCL, diffuse large B cell lymphoma CMML, Chronic Myelomonocytic Leukemia PCR+, monoclonal TCR rearrangement AITL, angioimmunoblastic T-cell lymphoma PTCL, peripheral T-cell lymphoma CTCL, cutaneous T-cell lymphoma MF, myelofibrosis MCF, multiple color flow cytometry

**Supplementary Table 2.**
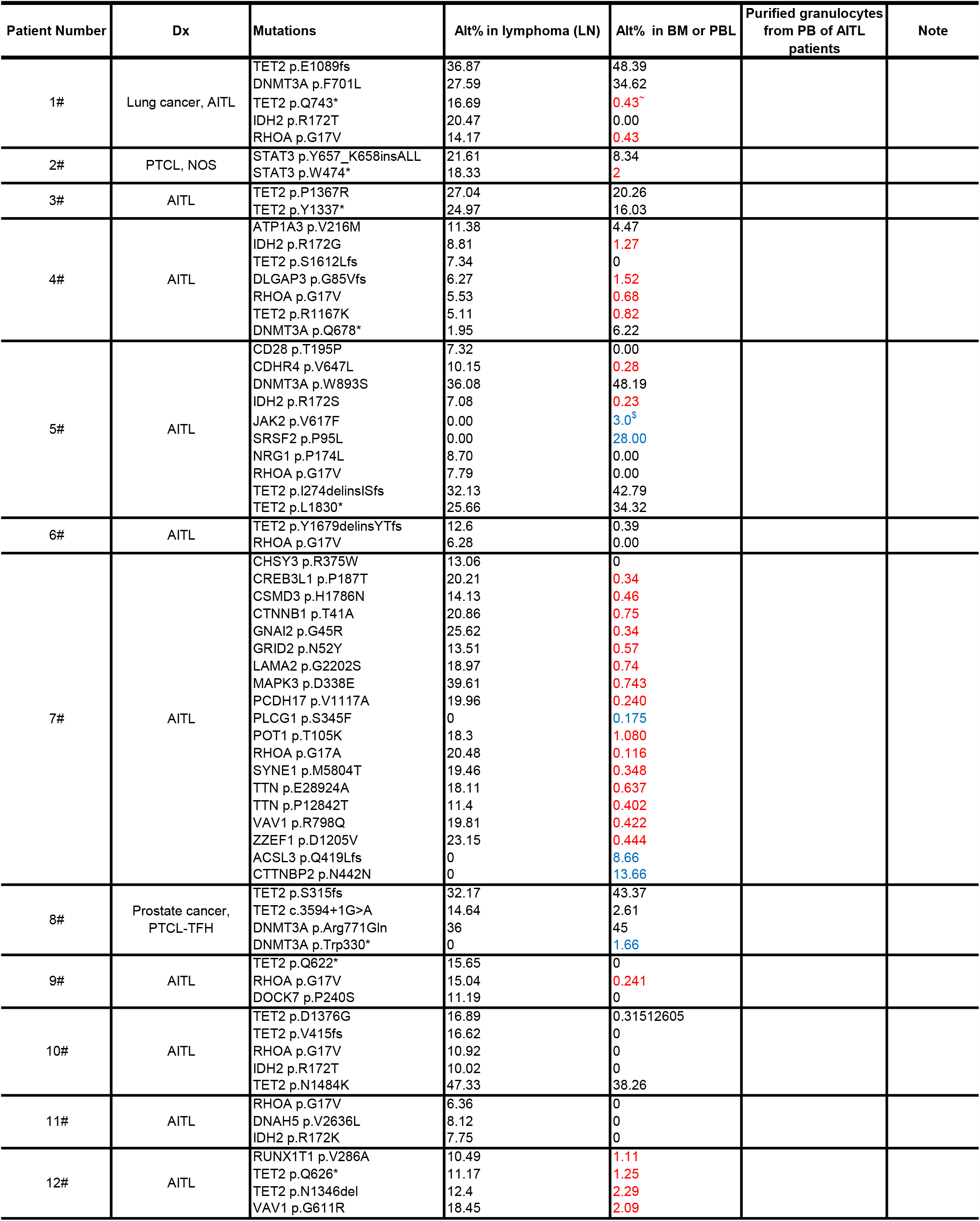
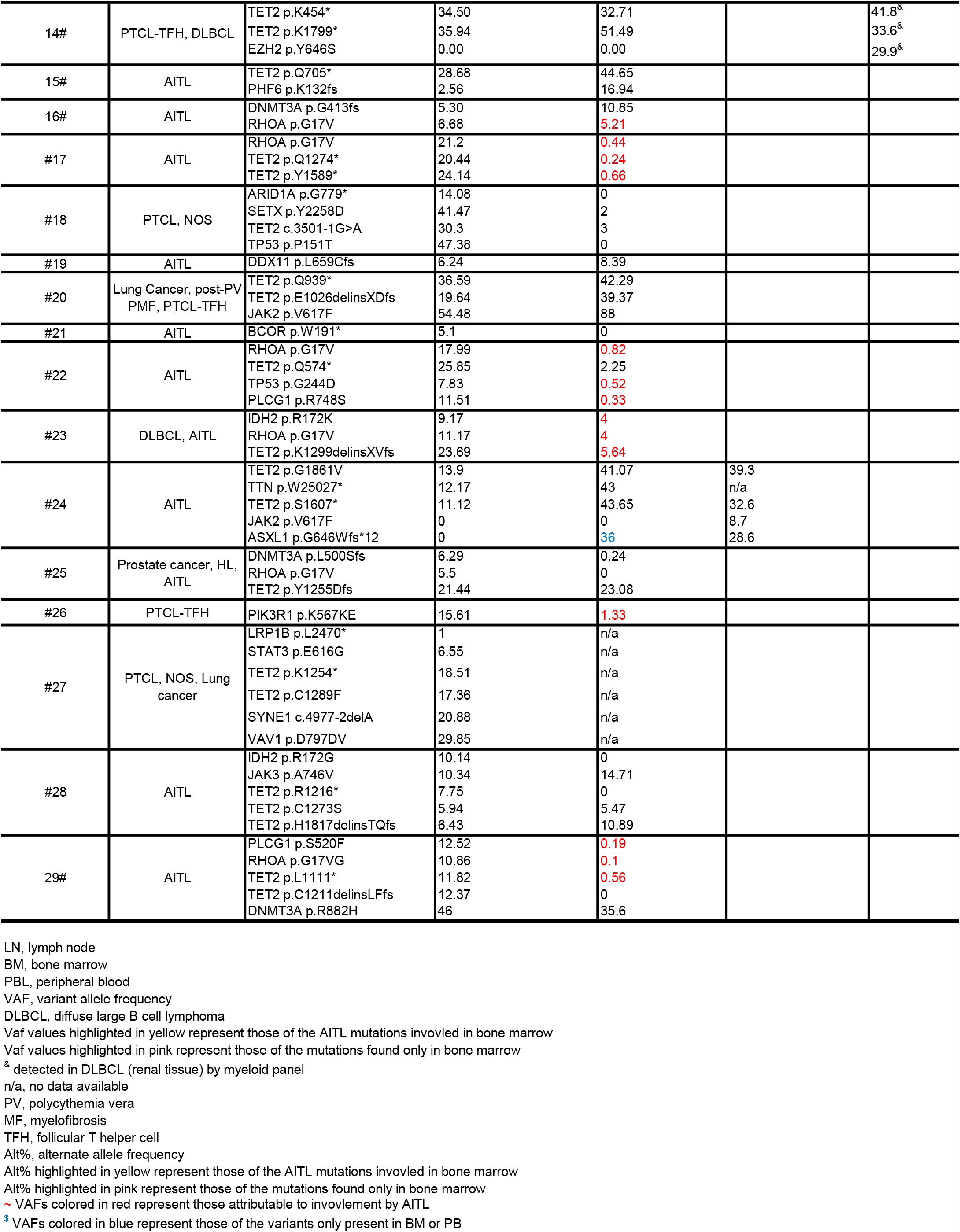
Individival mutations and their VAFs in the lymphoma cases and matched BM/PB.

**Supplementary Table 3.** Variant summary.

|  |  | Primary AITL lymphoma |  |  |  | Paired BM/PB<sup>§</sup> |  |  |  |
| --- | --- | --- | --- | --- | --- | --- | --- | --- | --- |
|  |  | summary | Mean | Median | Percent | summary | Mean | Median | Percent |
| Variant Classification | Frame_Shift_Del | 9 | 0.333 | 0 | 8.82 | 6 | 0.316 | 0 | 13.64 |
|  | Frame_Shift_Ins | 7 | 0.259 | 0 | 6.86 | 7 | 0.316 | 0 | 15.91 |
|  | In_Frame_Del | 1 | 0.037 | 0 | 0.98 | 0 | 0 | 0 | 0.00 |
|  | In_Frame_Ins | 1 | 0.037 | 0 | 0.98 | 1 | 0.053 | 0 | 2.27 |
|  | Missense_Mutation | 62 | 2.296 | 1 | 60.78 | 17 | 0.789 | 1 | 38.64 |
|  | Nonsense_Mutation | 20 | 0.741 | 1 | 19.61 | 12 | 0.579 | 0 | 27.27 |
|  | Splice_Site | 2 | 0.074 | 0 | 1.96 | 1 | 0.053 | 0 | 2.27 |
| Total |  | 102 | 3.778 | 3 | 100.00 | 44 | 2.105 | 2 | 100.00 |
| No. of Samples |  | 27 |  |  |  | 27* |  |  |  |
| No. of Mutated genes |  | 37 |  |  |  | 14 |  |  |  |

\*A total of 23 bone marrow and 4 PBL samples were sequenced. Among 27 BM/PB samples, 20 had identifiable mutated genes after filtering out the variants involved.
<sup>§</sup> the variants from the invovled lymphoma excluded

## Notes

### Competing Interest Statement

The authors have declared no competing interest.

### Funding Statement

This study is supported by a grant (R01 CA194547) from the National Cancer Institute to Dr. Wayne Tam.

### Author Declarations

the Institutional Review Board of Weill Cornell Medicine, New York, USA

